# Surge capacities and predicted demands of Brazil’s health system associated with severe COVID-19 cases

**DOI:** 10.1101/2020.04.02.20050351

**Authors:** Eric Luis Barroso Cavalcante, Juliana Cristina Cardoso Ferreira

**Author notes:** Corresponding author. *E-mail address:* (Eric Luis Barroso Cavalcante).

## Abstract

**Objective:** To estimate surge capacities and demands of Brazil’s health system in view of severe cases of the novel coronavirus disease.

**Methods:** Three types of hospital equipments demanded by severe COVID-19 patients are considered: available intensive care unit (ICU) beds, existing surgery operating rooms and respirators located in other hospital areas. They are taken into account on a cumulative basis forming three levels of hospital equipment usage. Based on a mean duration of hospitalization for the disease, it is estimated the daily admission capacity of infected patients per state and for the entire country for each level of hospital equipment usage. Furthermore, an exponential regression model is fitted by means of the daily national number of new documented patients. The prediction intervals for the number of new patients for certain days in the immediate future are then calculated and compared to the admissible daily demand for the three incremental groups of equipments. The data are made publicly available by the Brazilian federal government and are gathered and analyzed by means of the Python programming language.

**Results:** 41% of the (adult) ICU beds in Brazil were available during 2019, indicating that this hospital equipment has not been on average operating near capacity in national numbers. Nevertheless, there is a marked heterogeneity in the absolute and relative numbers of available and existing ICU beds and existing surgery operating rooms and extra respirators between its states.

**Conclusions:** The remarkable differences between states’ hospital resources directly reflect into the number of possible daily admissions of COVID-19 patients with respiratory failure for the three considered levels of hospital equipment usage. In national numbers, Brazil’s health system is estimated to be capable of daily admitting 693, 1243 and 2166 severe patients under the three studied scenarios. The fitted model predicted that only the first limit, if any, would be reached.

## Introduction

Starting in the state capital of Wuhan, China, in December 2019, the novel coronavirus (SARS-COV-2) has been spreading rapidly all over the world and the World Health Organization (WHO) declared its disease, COVID-19, as a pandemic on 11th March 2020. One and a half month later the number of confirmed infections neared the three millions mark and 200000 deaths and reached almost every country and territory across the world, according to the Johns Hopkins University tracker (https://www.jhu.edu/).

Like the pandemic influenza in 1918, COVID-19 is associated with respiratory spread, an undetermined percentage of infected people with presymptomatic or asymptomatic cases transmitting infection to others, and a high fatality rate^1^.

Ji *et al*., after performing analysis of 70641 confirmed cases and 1772 deaths due to COVID-19 and assuming that average levels of healthcare are similar throughout China, consider that higher numbers of infections in a given population can be considered an indirect indicator of a heavier healthcare burden. By plotting the mortality rate against the incidence of the disease (cumulative number of confirmed cases since the start of the outbreak, per 10000 population) they have found a significant positive correlation, suggesting that mortality is correlated with healthcare burden and, consequently, with healthcare resource availability in an inverse correlation^2^.

In this fashion, the present work mainly proposes to estimate surge capacities of Brazil’s health system associated with severe cases of COVID-19. Three incremental levels of hospital equipment usage are considered: (1) in terms of available intensive care unit (ICU) beds; (2) in terms of available ICU beds and existing surgery operating rooms; and (3) in terms of available ICU beds and existing surgery operating rooms and respirators located in other hospital areas. Based on a reference mean duration of hospitalization for the disease, it is estimated the daily admission capacity of infected patients per state and for the entire country for each level of hospital equipment usage.

To the best of the authors’ knowledge it is the first public attempt to estimate (without mathematical simulation) the average availability of critical care beds in hospitals of the entire country deducting from the existing beds the equivalent number of beds that has been occupied during 2019 on a monthly average^3,4^. Two even more recent studies than the ones performed by Moraes^3^ and Goldwasser *et al*.^4^ also do not address the occupancy of ICU beds^5,6^.

Moreover, an exponential regression model is fitted using the available data for the daily national number of new patients who have been documented as infected. The prediction intervals for the number of new patients for certain days in the immediate future are then calculated and compared to the admissible daily demand for the three incremental groups of equipments.

## Modeling capacities

The data sources for estimating the hospital equipments available to treat severe COVID-19 patients are two health information systems: the National Registry of Health Facilities of Brazil (CNES) and the Hospital Information System of the Brazilian Public Health System (SIHSUS). The CNES data, obtained per facility, concern: (1) the number of existing ICU beds dedicated to the Brazilian Public Health System (SUS) (*x*_existing public ICUb_) and the number of existing ICU beds private to a for-profit healthcare provider (*x*_existing private ICUb_);(1) the number of existing operating rooms dedicated to surgeries (*x*_oper. rooms_); and (3) the number of existing respirators (*x*_resps._). The SIHSUS data, aggregated per facility, concern the number of ICU-days-per-month of beds dedicated to SUS (*x*_public ICU-days/month_). These data are available in the File Transfer Protocol server’s internet address (ftp://ftp.datasus.gov.br/) of the Brazilian Health Informatics Department (DATASUS) and have been downloaded and gathered in a local (personal computer) database by means of a Python programming package developed by one of the authors and currently available on the internet at https://github.com/SecexSaudeTCU/Pegasus/tree/master/pegasus/dados.

The types of ICU beds taken into account are the ones that are designed for adults, including the ones for burned, coronary and isolated patients and excluding the neonatal and pediatric beds^6^. It has been taken into account only the healthcare facilities that have at least one ICU bed dedicated to SUS and that remained as such during all months of 2019. Each one of the five healthcare facility quantities considered and mentioned on the previous paragraph have been arithmetically averaged through the months of 2019, leading to:

- 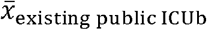: monthly mean of existing public ICU beds;
- 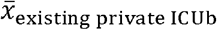: monthly mean of existing private ICU beds;
- 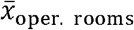: monthly mean of existing surgery operating rooms;
- 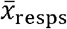: monthly mean of existing respirators;
- 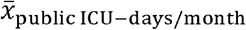: monthly mean of ICU-days-per-month of beds dedicated to SUS.

First, it is computed the monthly mean of existing ICU beds (for each healthcare facility):

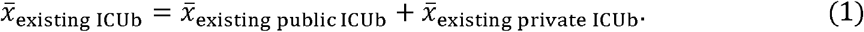

One can also write the vector form of (1):

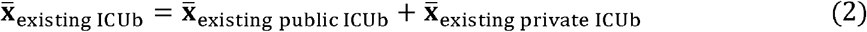

with

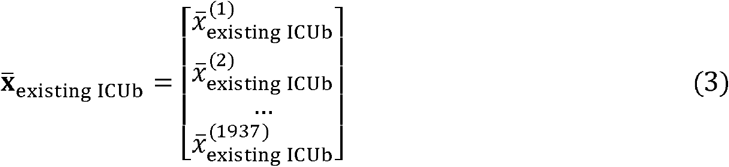

where 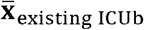 represents the vector of monthly mean existing ICU beds for the 1937 healthcare facilities that comply with the aforementioned criteria. In fact, all of the average quantities defined previously can be expressed in this compacted vector form that aggregates all the 1937 healthcare facilities. For conciseness, in the sequel we shall omit “monthly mean” from the expression “vector of monthly mean”.

To compute the vector of available public ICU beds the following expression is utilized:

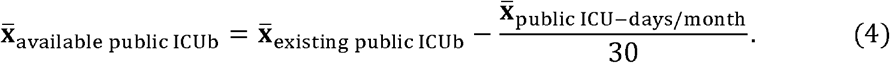

The vector of available private ICU beds is calculated considering the same proportion of public ICU bed usage:

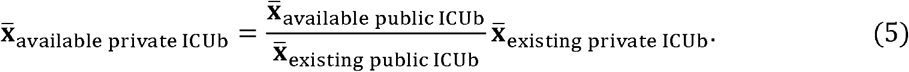

Now, one can compute the vector of available ICU beds:

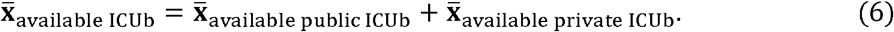

Also, the respirators that surpass the ones present in existing ICU beds and in surgery operating rooms are named extra respirators and computed accordingly to^7^:

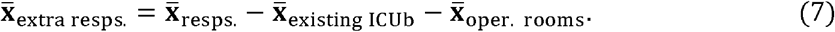

The vectors 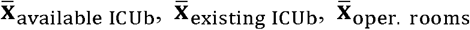. and 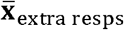. are dealt with in the “Results” section.

## Modeling predictions

The daily number of documented COVID-19 infections in Brazil between 26th February 2020 and 22th April 2020 is provided by DATASUS at https://covid.saude.gov.br/, and it has allowed to fit, using ordinary least squares, a two-degree exponential regression model to represent the trained part of this dataset comprised in total of 51 observations:

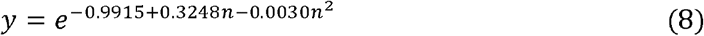

where *n* is the *n*th day since 26th February 2020 and *y* is the number of new COVID-19 cases on the *n*th day. Equation (8) represents the best fit among one to eight-degree exponential regression models and one to eight-degree polynomial regression models using the *R*^*2*^ score as statistical measure. Each model has been trained on 70% of the randomly chosen observations and the test *R*^*2*^ score is 0·98 and it has been computed on the 30% remaining observations.

## Results

The scatter plot of the daily new number of COVID-19 cases and the curve of the best model (8) are illustrated in Figure 1. In an informative capacity, it is also shown the scatter plot of the cumulative number of COVID-19 cases against the date in Figure 2.

**Figure 1.**
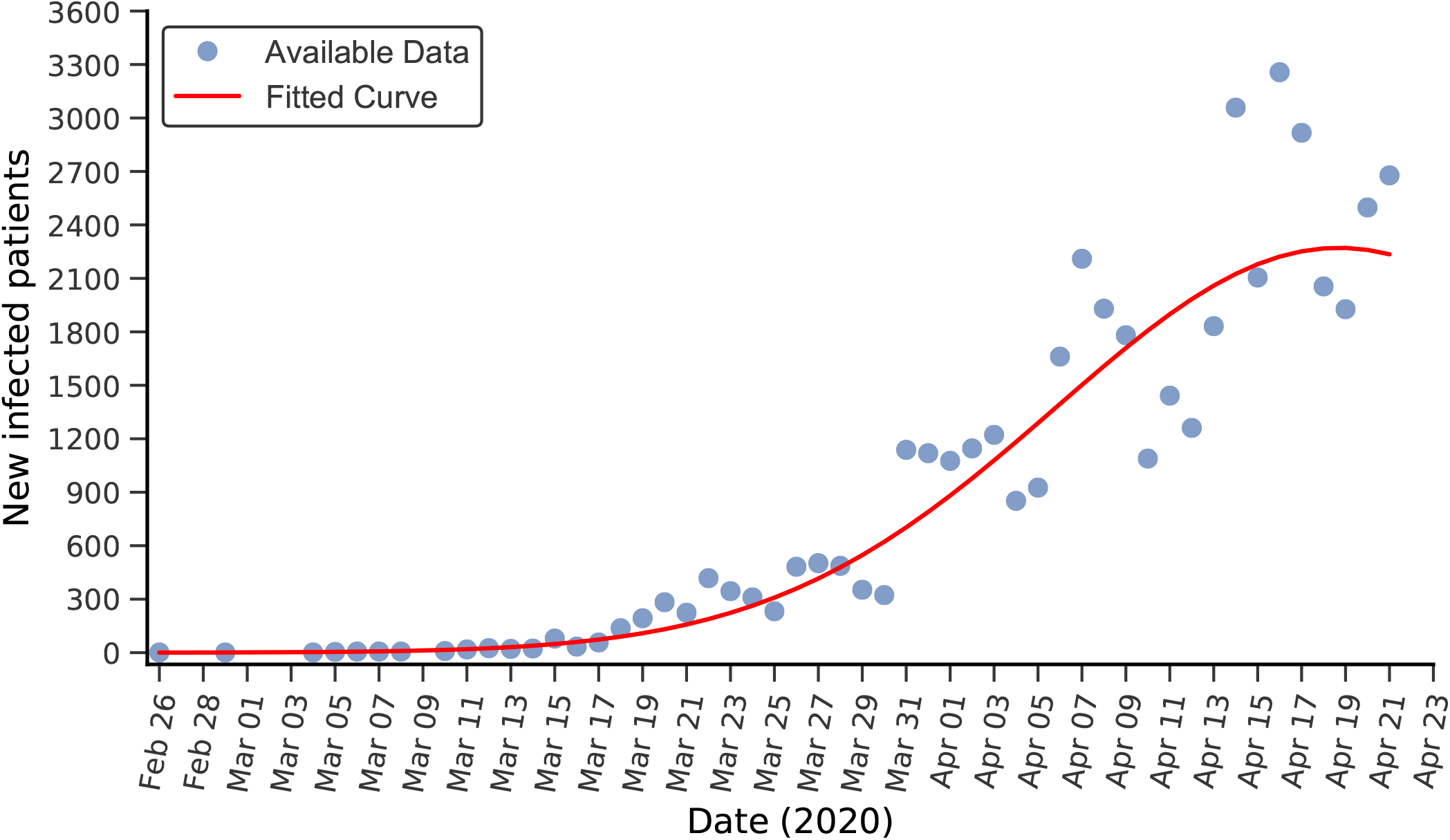
Measured and predicted new number of documented COVID-19 infected patients in Brazil

**Figure 2.**
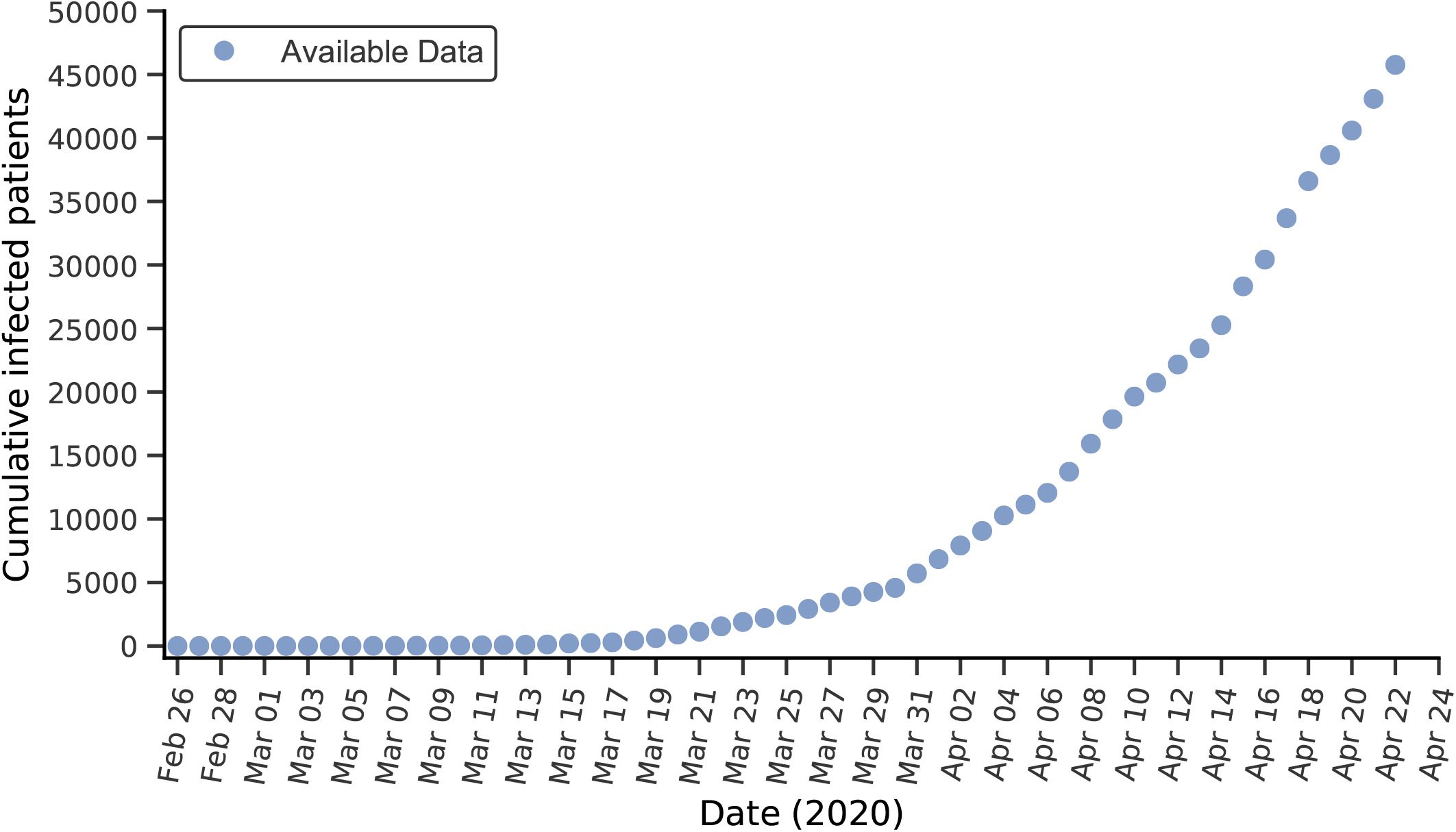
Measured cumulative number of documented COVID-19 infected patients in Brazil

Four vectors (of size 1937) that represent quantities of hospital equipments and are defined in the “Modeling capacities” section are vertically aligned and gathered as Table 1, which thus contains: the monthly mean of available and existing (adult) ICU beds (excluding neonatal and pediatric units), the monthly mean of existing surgery operating rooms and the monthly mean of extra respirators per healthcare facility (only listed six facilities). It can be seen that, during the year of 2019, 9021 ICU beds were on average available in Brazil out of 21908 that are appropriate to accommodate adult COVID-19 patients. It is as if at any given instant of 2019 41% of the ICU beds in the country were available, indicating that this hospital equipment has not been on average operating near or at capacity in terms of national numbers and it is worth mentioning that this ICU availability rate is 7·6 percentage points larger than in the United States^7^. Table 2 aggregates the data of Table 1 per state (including the federal capital territory Distrito Federal (DF)).

**Table 1.**
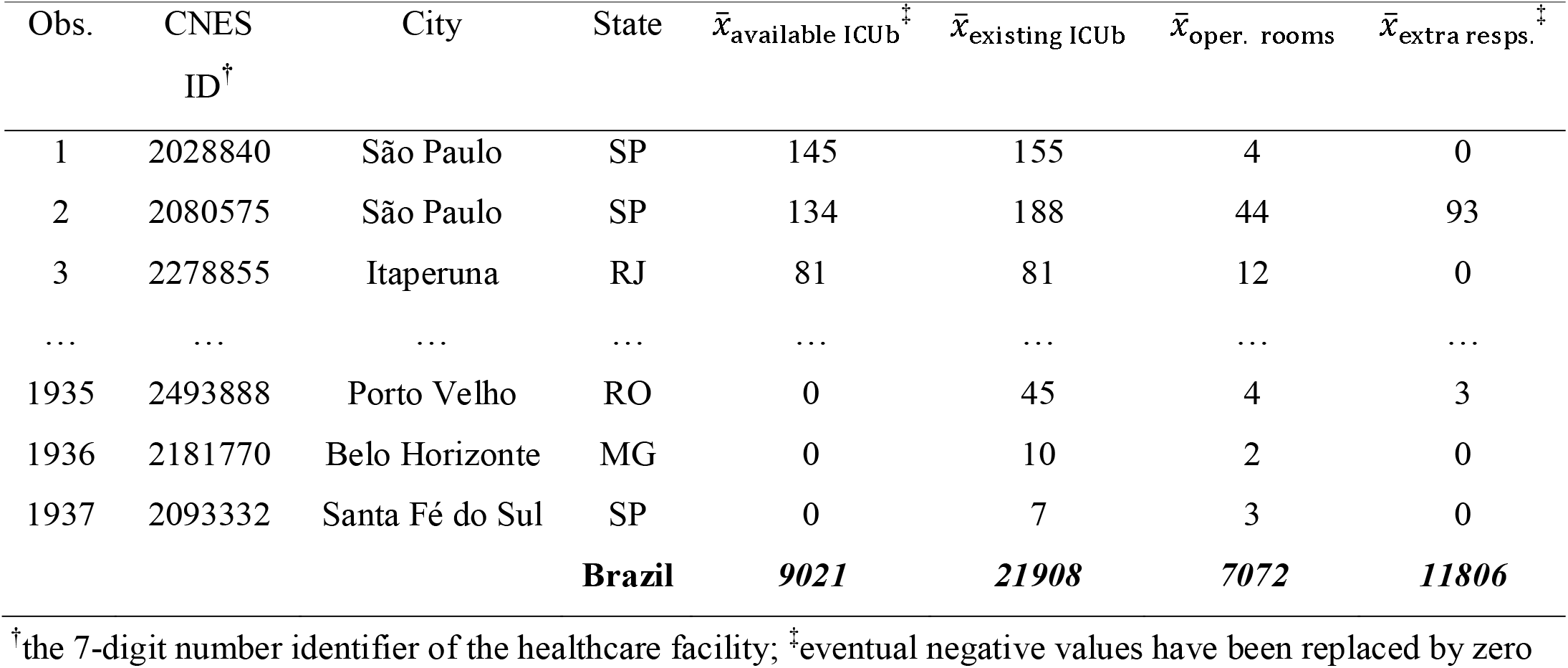
Mean of available and existing ICU beds, existing surgery oper. rooms and extra respirators

**Table 2.** Mean of available and existing ICU beds, existing surgery oper. rooms and extra respirators per state

| State | $\bar{x}_1$ | $\bar{x}_2$ | $\bar{x}_3$ | $\bar{x}_4$ | State | $\bar{x}_1$ | $\bar{x}_2$ | $\bar{x}_3$ | $\bar{x}_4$ |
| --- | --- | --- | --- | --- | --- | --- | --- | --- | --- |
| AP | 28 | 52 | 21 | 0 | AM | 207 | 316 | 107 | 241 |
| AC | 29 | 55 | 21 | 45 | MS | 65 | 252 | 91 | 264 |
| TO | 102 | 101 | 42 | 49 | ES | 205 | 587 | 148 | 300 |
| PI | 134 | 233 | 116 | 79 | DF | 144 | 324 | 74 | 305 |
| RR | 20 | 40 | 18 | 80 | SC | 294 | 769 | 289 | 383 |
| RO | 51 | 189 | 41 | 85 | PR | 464 | 1787 | 517 | 403 |
| AL | 131 | 255 | 88 | 124 | CE | 384 | 648 | 225 | 465 |
| SE | 122 | 221 | 52 | 131 | BA | 655 | 1254 | 378 | 488 |
| RN | 133 | 334 | 94 | 145 | PE | 471 | 1127 | 263 | 604 |
| MA | 226 | 445 | 194 | 153 | RS | 600 | 1504 | 549 | 702 |
| GO | 388 | 777 | 235 | 162 | MG | 763 | 2672 | 841 | 960 |
| PB | 238 | 445 | 159 | 193 | RJ | 849 | 1768 | 593 | 1256 |
| MT | 170 | 352 | 143 | 220 | SP | 1670 | 4838 | 1551 | 3745 |
| PA | 478 | 563 | 222 | 224 | **Brazil** | **9021** | **21908** | **7072** | **11806** |

$\bar{x}_1$ : $\sum \bar{x}_{\text{available ICUb}}$ ; $\bar{x}_2$ : $\sum \bar{x}_{\text{existing ICUb}}$ ; $\bar{x}_3$ : $\sum \bar{x}_{\text{oper. rooms}}$ ; $\bar{x}_4$ : $\sum \bar{x}_{\text{extra resps.}}$

Table 3 depicts the equivalent quantities of Table 2 in terms of 10000 inhabitants. The 2019 population data that allow calculate these rates are present in the website of the Brazilian Institute of Geography and Statistics (IBGE) (https://www.ibge.gov.br/). Comparing Table 2 and Table 3 it can be seen that the Roraima state (RR) is the most precarious in terms of available and existing ICU beds and existing operating rooms (20; 40; 18), and the most precarious in terms of available ICU beds per 10000 inhabitants is the state of Mato Grosso do Sul (MS) with 0·23. On the other hand, the São Paulo (SP) state has the largest absolute number of existing ICU beds (4838), but the state of Tocantins (TO) appears as the most equipped in terms of available ICU beds per 10000 (0·65). The Brazilian national rate of existing (adult) ICU beds per 10000 (1·04) is less than a third of the United States rate in the year of 2009 (3·47)^8^.

**Table 3.**
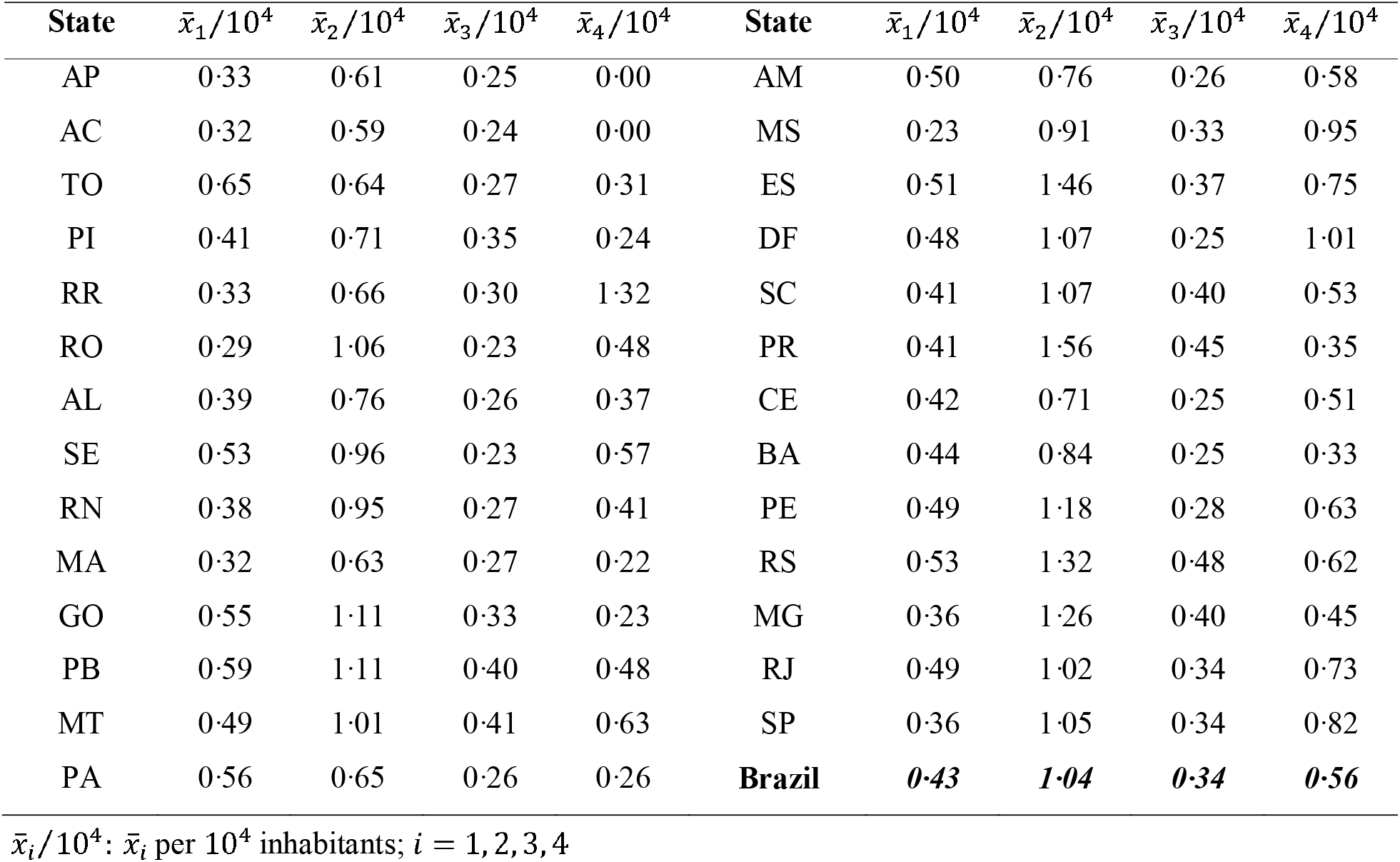
Rate of available and existing ICU beds, existing surgery oper. rooms and extra respirators per state

| State | $\bar{x}_1/10^4$ | $\bar{x}_2/10^4$ | $\bar{x}_3/10^4$ | $\bar{x}_4/10^4$ | State | $\bar{x}_1/10^4$ | $\bar{x}_2/10^4$ | $\bar{x}_3/10^4$ | $\bar{x}_4/10^4$ |
| --- | --- | --- | --- | --- | --- | --- | --- | --- | --- |
| AP | 0.33 | 0.61 | 0.25 | 0.00 | AM | 0.50 | 0.76 | 0.26 | 0.58 |
| AC | 0.32 | 0.59 | 0.24 | 0.00 | MS | 0.23 | 0.91 | 0.33 | 0.95 |
| TO | 0.65 | 0.64 | 0.27 | 0.31 | ES | 0.51 | 1.46 | 0.37 | 0.75 |
| PI | 0.41 | 0.71 | 0.35 | 0.24 | DF | 0.48 | 1.07 | 0.25 | 1.01 |
| RR | 0.33 | 0.66 | 0.30 | 1.32 | SC | 0.41 | 1.07 | 0.40 | 0.53 |
| RO | 0.29 | 1.06 | 0.23 | 0.48 | PR | 0.41 | 1.56 | 0.45 | 0.35 |
| AL | 0.39 | 0.76 | 0.26 | 0.37 | CE | 0.42 | 0.71 | 0.25 | 0.51 |
| SE | 0.53 | 0.96 | 0.23 | 0.57 | BA | 0.44 | 0.84 | 0.25 | 0.33 |
| RN | 0.38 | 0.95 | 0.27 | 0.41 | PE | 0.49 | 1.18 | 0.28 | 0.63 |
| MA | 0.32 | 0.63 | 0.27 | 0.22 | RS | 0.53 | 1.32 | 0.48 | 0.62 |
| GO | 0.55 | 1.11 | 0.33 | 0.23 | MG | 0.36 | 1.26 | 0.40 | 0.45 |
| PB | 0.59 | 1.11 | 0.40 | 0.48 | RJ | 0.49 | 1.02 | 0.34 | 0.73 |
| MT | 0.49 | 1.01 | 0.41 | 0.63 | SP | 0.36 | 1.05 | 0.34 | 0.82 |
| PA | 0.56 | 0.65 | 0.26 | 0.26 | **Brazil** | **0.43** | **1.04** | **0.34** | **0.56** |

$\bar{x}_i/10^4$ : $\bar{x}_i$ per $10^4$ inhabitants; $i = 1, 2, 3, 4$

Table 4, also constructed based on Table 2, illustrates the available capacity of Brazil’s health system to face severe COVID-19 cases considering three incremental levels of hospital equipment usage: (1) in terms of available ICU beds (named Level 1; 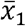); (2) available ICU beds and existing surgery operating rooms (Level 2; 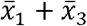); and (3) available ICU beds and existing surgery operating rooms and respirators located in other hospital areas (Level 3; 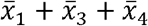). It is thus assumed that all existing surgery operating rooms and respirators located in other hospital areas (computed accordingly to equation (7)) can be allocated to attend the critically infected by COVID-19.

**Table 4.**
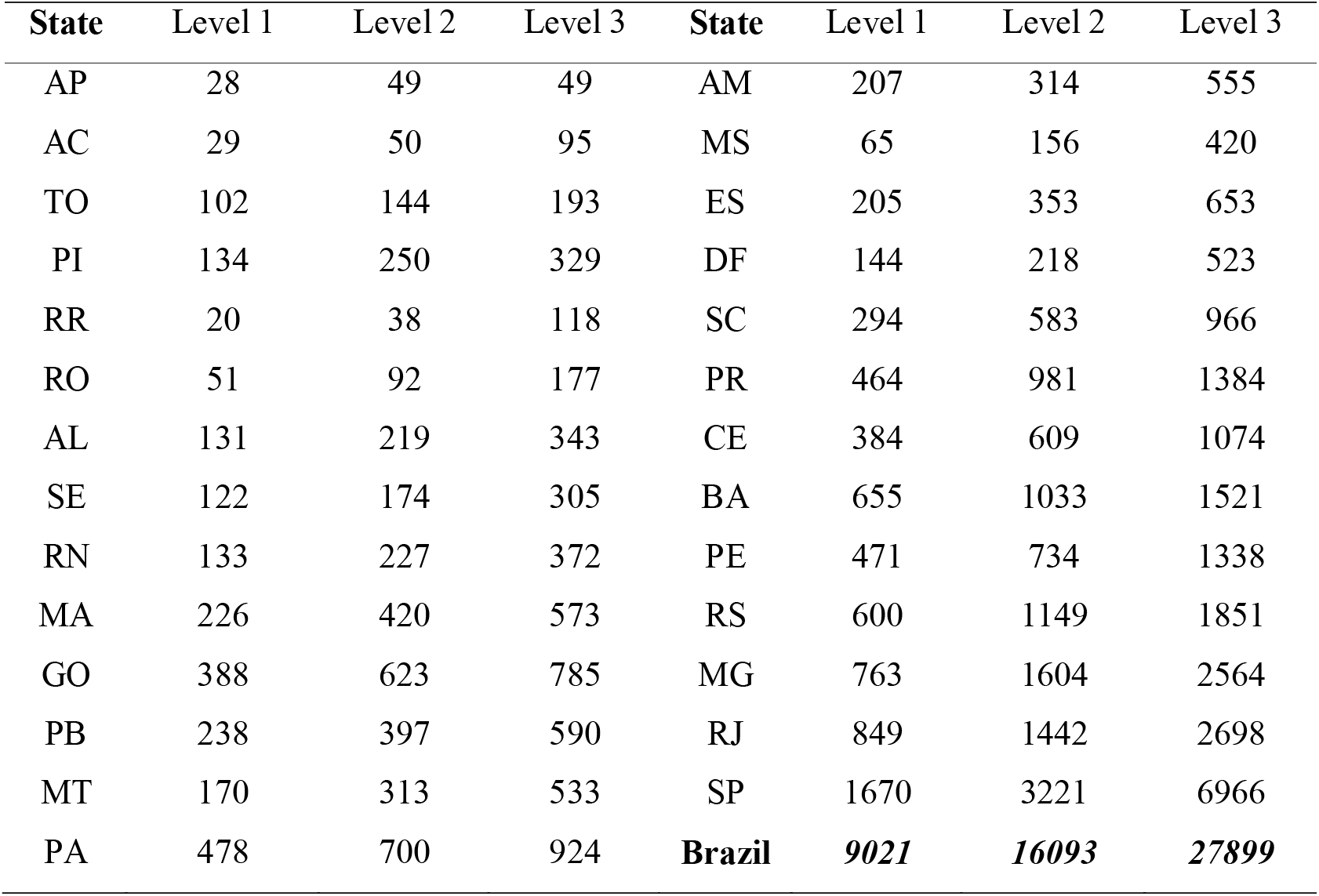
Capacities of Brazil’s health system to face severe COVID-19 cases per state

| State | Level 1 | Level 2 | Level 3 | State | Level 1 | Level 2 | Level 3 |
| --- | --- | --- | --- | --- | --- | --- | --- |
| AP | 28 | 49 | 49 | AM | 207 | 314 | 555 |
| AC | 29 | 50 | 95 | MS | 65 | 156 | 420 |
| TO | 102 | 144 | 193 | ES | 205 | 353 | 653 |
| PI | 134 | 250 | 329 | DF | 144 | 218 | 523 |
| RR | 20 | 38 | 118 | SC | 294 | 583 | 966 |
| RO | 51 | 92 | 177 | PR | 464 | 981 | 1384 |
| AL | 131 | 219 | 343 | CE | 384 | 609 | 1074 |
| SE | 122 | 174 | 305 | BA | 655 | 1033 | 1521 |
| RN | 133 | 227 | 372 | PE | 471 | 734 | 1338 |
| MA | 226 | 420 | 573 | RS | 600 | 1149 | 1851 |
| GO | 388 | 623 | 785 | MG | 763 | 1604 | 2564 |
| PB | 238 | 397 | 590 | RJ | 849 | 1442 | 2698 |
| MT | 170 | 313 | 533 | SP | 1670 | 3221 | 6966 |
| PA | 478 | 700 | 924 | **Brazil** | **9021** | **16093** | **27899** |

Assuming the mean duration of hospitalization as 12·8 days, one can estimate the possible daily admission of patients with respiratory failure for each of the three incremental groups of hospital equipments considered in Table 4^9^. Such rates are shown in Table 5 as Scenario 1, Scenario 2 and Scenario 3, respectively. The Brazilian state that can admit the least number of severe COVID-19 patients considering only the usage of available ICU beds is RR (1 per day) and, in the worst-case scenario (demand of the available ICU beds and existing operating rooms and respirators located in other hospital areas), the state with the worst number is Amapá (AP): 3 possible admissions per day. On the other extreme, it is the state of SP, which can daily admit the largest number of severe patients for all scenarios (130, 251, 544), but also is the most populated state, and has the biggest Italian community of Brazil, the most internationally connected Brazilian airport and the two most busiest Brazilian airports in terms of domestic passenger traffic^10-12^. It is worth noting that, at least up until 22th April 2020, Italy was one of the three countries most impacted by COVID-19 and the SP state had the fifth largest number of documented cases of the coronavirus disease per inhabitant.

**Table 5.** Daily admission number of patients with respiratory failure per state

| **State** | Scenario 1 | Scenario 2 | Scenario 3 | **State** | Scenario 1 | Scenario 2 | Scenario 3 |
| --- | --- | --- | --- | --- | --- | --- | --- |
| AP | 2 | 3 | 3 | AM | 16 | 24 | 43 |
| AC | 2 | 3 | 7 | MS | 5 | 12 | 32 |
| TO | 7 | 11 | 15 | ES | 16 | 27 | 51 |
| PI | 10 | 19 | 25 | DF | 11 | 17 | 40 |
| RR | 1 | 2 | 9 | SC | 22 | 45 | 75 |
| RO | 3 | 7 | 13 | PR | 36 | 76 | 108 |
| AL | 10 | 17 | 26 | CE | 30 | 47 | 83 |
| SE | 9 | 13 | 23 | BA | 51 | 80 | 118 |
| RN | 10 | 17 | 29 | PE | 36 | 57 | 104 |
| MA | 17 | 32 | 44 | RS | 46 | 89 | 144 |
| GO | 30 | 48 | 61 | MG | 59 | 125 | 200 |
| PB | 18 | 31 | 46 | RJ | 66 | 112 | 210 |
| MT | 13 | 24 | 41 | SP | 130 | 251 | 544 |
| PA | 37 | 54 | 72 | **Brazil** | **693** | **1243** | **2166** |

It can be argued from Table 5 that if it reaches a point at which Brazil’s health system is daily admitting more than 693, 1243 and 2166 patients with respiratory failure the system may demonstrate stress under Scenario 1, Scenario 2 and Scenario 3, respectively. In this way, assuming that 15% of documented COVID-19 infections result in severe disease the daily numbers of new COVID-19 cases would have to be at maximum 4620 (693/0.15), 8287 (1243/0.15) and 14440 (2166/0.15) in order to not demand Brazil’ health system beyond the limits of Scenario 1, Scenario 2 and Scenario 3, respectively^13^.

According to the fitted exponential regression model (8), the lower bound values of its 90% predictions intervals indicate that the admissible numbers of daily new infected people of Scenario 1 (4620), Scenario 2 (8287) or Scenario 3 (14440) would not be reached in April 2020. Conversely, the upper bounds revel that only the limit of Scenario 1, consisted of available ICU beds, would be first reached on the 14th of April 2020. The 90% prediction interval for this date is [915, 4633], which means that 90% of intervals of these forms will contain the true number of new COVID-19 cases for this date. Along with the uncertainty that resides in the own definition of prediction intervals, their ranges for the fitted model are quite significant, expressing another source of uncertainty regarding the true number of new COVID-19 cases for a certain date.

## Discussions

Acknowledging, as suggested by Ji *et al*., the potential negative association of COVID-19 mortality rate with healthcare resource availability might help Brazil to be better prepared for its outbreak^2^. In this way, it has been estimated that 41% of the (adult) ICU beds in Brazil were available during 2019, indicating that this hospital equipment is not on average operating near or at capacity in terms of national numbers and it is worth mentioning that this ICU availability rate is 7·6 percentage points larger than in the United States^7^. On the other hand, the Brazilian rate of existing (adult) ICU beds per 10000 (1·04) is less than a third of the United States rate in the year of 2009 (3·47)^8^.

Moreover, there is a marked heterogeneity in the absolute and relative numbers of available and existing ICU beds and existing surgery operating rooms and extra respirators between the states of Brazil (including its federal capital territory). The remarkable differences between states’ hospital resources directly reflect into the number of possible admissions per day of COVID-19 patients with respiratory failure for each of the three levels of hospital equipment usage.

In national numbers, Brazil’s health system is estimated to be capable of daily admitting at maximum 693, 1243 and 2166 patients with respiratory failure under the three studied scenarios and the fitted exponential growth model indicates that Brazil’ health system will not reach the limit of Scenario 2 and Scenario 3 in April and might or not reach the limit of Scenario 1 on April 2020.

This work concern data of 2019 and, consequently, takes into account only hospital equipments available previously to the COVID-19 disease outbreak. Additionally, it does not include hospitals that do not have at least one critical care bed dedicated to SUS.

## Data Availability

All data referred to in the manuscript are available on the internet for public downloading.

ftp://ftp.datasus.gov.br/dissemin/publicos/

https://www.ibge.gov.br/estatisticas/sociais/populacao/9103-estimativas-de-populacao.html?=&t=downloads

https://covid.saude.gov.br/

## Contributors

ELBC was responsible for literature search, data collection, data analysis, statistics, data interpretation and writing of the manuscript. JCCF was responsible for literature search, data interpretation and writing of the manuscript.

## References

1 Morens DM, Daszak P, Taubenberger JK. Escaping Pandora’s box - another novel coronavirus. The New England Journal of Medicine 2020; published online February 26.

2 Ji Y, Ma Z, Peppelenbosch MP, Pan Q. Potential association between COVID-19 mortality and health-care resource availability. Lancet Global Health 2020; published online February 25.

3 Moraes RS. Modeling and simulation of public ICU bed usage in the state of Rio de Janeiro (in Portuguese). Master’s Dissertation in Production Engineering 2014; Federal University of Rio de Janeiro, Rio de Janeiro.

4 Goldwasser RS, Lobo MSC, Arruda EF, et al. Difficulties in access and estimates of public beds in intensive care units in the state of Rio de Janeiro. Rev Saúde Pública 2016; 50: 19–19.

5 Brazilian Federal Council of Medicine. Less than 10% of the Brazilian counties possess ICU beds. Available from https://portal.cfm.org.br/index.php?option=com_content&view=article&id=27828:2018-09-04-19-31-41&catid=3. Accessed in April 5, 2020.

6 Department of Economics and Statistics of the Rio Grande do Sul (RS) state. Coronavirus RS scenario. Available from https://planejamento.rs.gov.br/cenariocoronavirus. Accessed in April 5, 2020.

7 Halpern NA, Tan KS. U.S. ICU resource availability for COVID-19. Society of Critical Care Medicine 2020; March 13. Available from https://sccm.org/Blog/March-2020/United-States-Resource-Availability-for-COVID-19. Accessed in April 5, 2020.

8 Wallace D J, Angus DC, Seymour CW, Barnato AE, Kahn JM. Critical care bed growth in the United States – A comparison of regional and national trends. American Journal of Respiratory and Critical Care Medicine 2015; 191 (4): 410–416.

9 Guan WJ, Ni ZY, Hu Y, et al. Clinical characteristics of coronavirus disease 2019 in China. The New England Journal of Medicine 2020; published online February 28.

10 Rosoli G. Italian Emigration to Brazil. Center for Migration Studies 1994; special issue 3 (11): 229–235.

11 OAG. Megahubs, Index 2019. Available from https://www.oag.com/oag-megahubs-2019. Accessed in April 5, 2020.

12 Couto GS, da Silva APC, Ruiz LB, Benevenuto F. Structural properties of the Brazilian air transportation network. Anais da Academia Brasileira de Ciências 2015; 87 (3): 1653–1674.

13 Cavallo JJ, Donoho DA, Forman HP. Hospital capacity and operations in the coronavirus disease 2019 (COVID-19) pandemic - planning for the nth patient. JAMA 2020; published online March 17.

